# Event-specific interventions to minimize COVID-19 transmission

**DOI:** 10.1101/2020.07.09.20149435

**Authors:** P. Tupper, H. Boury, M. Yerlanov, C. Colijn

## Abstract

Coronavirus disease 2019 (COVID-19) is a global pandemic with over 11 million cases worldwide. Currently there is no treatment and no vaccine. Interventions such as hand washing, masks, social distancing, and “social bubbles” are used to limit community transmission, but it is challenging to choose the best interventions for a given activity. Here, we provide a quantitative framework to determine which interventions are likely to have the most impact in which settings. We introduce the concept of “event R”, the expected number of new infections due to the presence of a single infected individual at an event. We obtain a fundamental relationship between event R and four parameters: transmission intensity, duration of exposure, the proximity of individuals, and the degree of mixing. We use reports of small outbreaks to establish event R and transmission intensity in a range of settings. We identify principles that guide whether physical distancing, masks and other barriers to transmission, or social bubbles will be most effective. We outline how this information can be obtained and used to re-open economies with principled measures to reduce COVID-19 transmission.

---

The global COVID-19 pandemic that began in late 2019 and spread rapidly around the world has been slowed by the widespread use of non-pharmaceutical interventions (NPIs), including border and travel restrictions, school closures, work-from-home edicts, the banning of mass gatherings and many other workplace and venue closures. These have been extremely costly economically, socially and for numerous health outcomes (*7*). Many jurisdictions are now moving to re-start economic and social activities, although unlike in past pandemics, they are doing so in the absence of meaningful levels of immunity to SARS-CoV-2. This leaves populations at high risk of resurgences in cases, and consequent high health care costs and lack of health care capacity.

Recent rises in COVID-19 cases in several US states and in countries around the world highlight the urgent need to understand how economic and social activity can be resumed while minimizing COVID-19 transmission risk. This remains unknown despite the pandemic’s large scale, with over 11M cases worldwide to date (*17*). Proposed actions that aim to reduce COVID-19 risk include face masks, plexiglass shields, pedestrian flow management, two metre distancing guidelines, reducing the capacity of many venues, and more. Many organizations must now make arrangements to re-open while attempting to reduce COVID-19 risk, in the near-complete absence of information about which measures will be most effective in their particular setting.

We have developed a conceptual framework and model to resolve some of the uncertainty around the effectiveness of different interventions. We build on the fundamental mathematical relationship between the number of people in contact with an infectious individual, the time for which they are in contact, and the risk of transmission per unit time. We inform our model with data from a set of reported events where transmissions occurred and were well characterized. To guide planners and provide an accessible framework, we focus on specific events and how transmission opportunities may differ under different interventions. We centre our discussion on what we call “event R”, or *eR*, namely the expected number of newly infected individuals *at an event* due to the attendance of a single infected individual.

Consider an event that lasts a total time *T*. If an infectious individual attends, and is in contact with a single susceptible individual for a time *τ* with a constant per unit time probability of transmission *β*, then the probability that the susceptible individual becomes infected is (1−*e*^−*βτ*^) (*27, Ch. 5*). (The constant rate assumption is a simplification that omits many factors that are known to be important; see Supplementary Information for discussion). If the infectious person contacts *k* others, instead of just one, then the expected number of new infections as a result of that contact is *k*(1 − *e*^−*βτ*^). Now suppose that instead of being in contact with the same group of *k* others, the event involves interacting with many groups of attendees. We model a simplified version of this type of mixing by imagining that for a time *τ*, the infectious attendee is in contact with *k* others, and then joins a new distinct group of *k* attendees for time *τ*, and so on. Over the course of the event, the infectious individual contacts on average *kT*/*τ* others, and the expected number of new infections that arises is

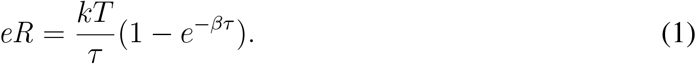

This fundamental equation relates the “event R” to the level of crowding at the event (which determines *k*), the level of mixing *T*/*τ* – do people contact mainly their “bubble” of nearby attendees or do they mix more widely – and the propensity for transmission in the physical setting (*β*).

In Fig. 1 (left) we illustrate three fundamentally different types of intervention that can be put in place to reduce the risk of COVID-19 transmission, and Eq. 1 allows us to examine when each will be most effective. Face masks, barriers, hand hygiene and similar measures aim to reduce the transmission rate *β*. Physical distancing measures that keep people apart reduce *k*. Finally, and less well-recognized, structuring the attendees into strict “social bubbles” and ensuring that people keep contact to within their bubble reduces mixing (increasing *τ*).

**Figure 1:**
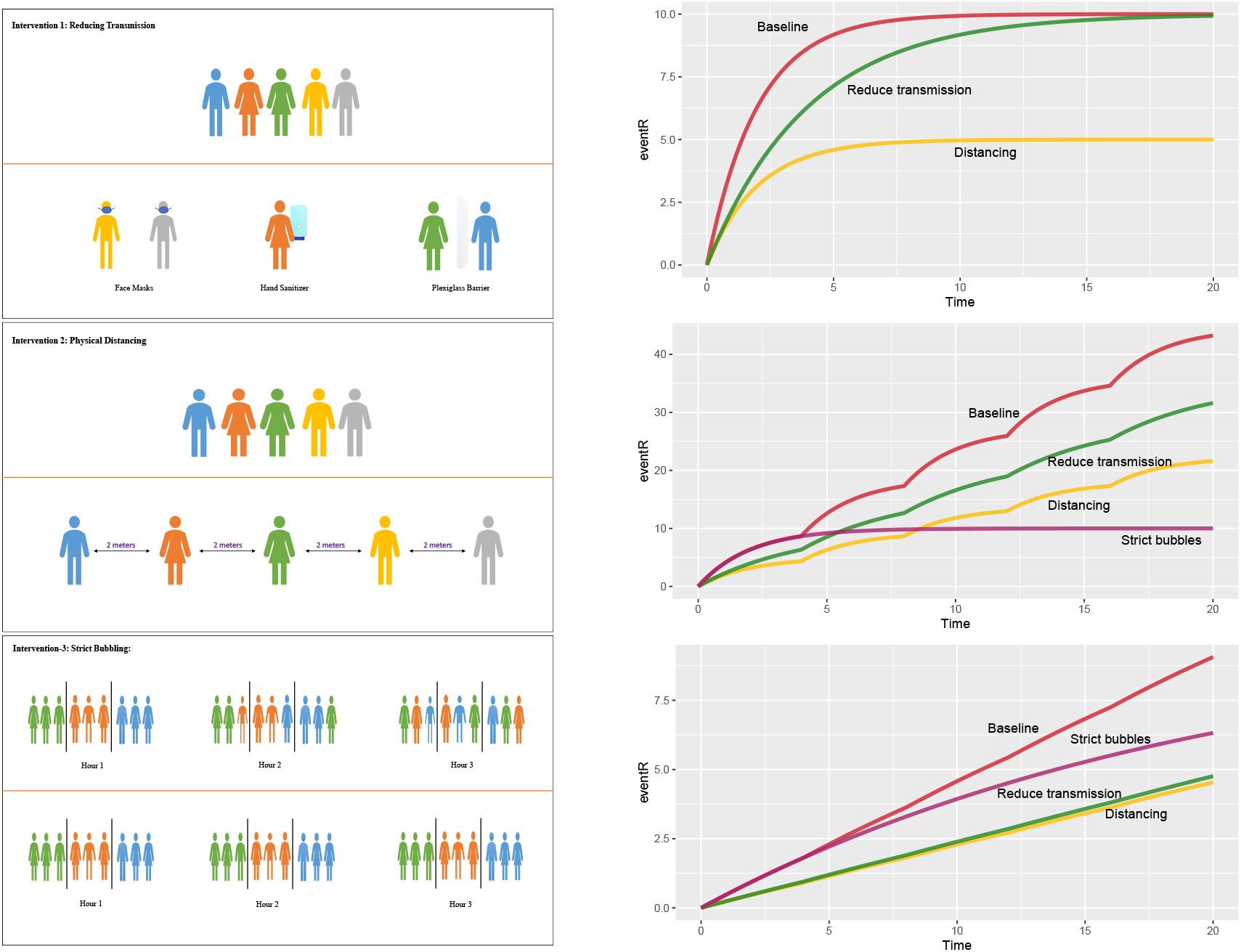
**Left**. The three types of intervention for reducing *eR* in a setting. *Top:* reducing transmission *β, Middle:* reducing the number of contacts at a given time *k, Bottom:* reducing mixing by increasing *τ*. **Right**. The effects of these interventions on *eR*. At baseline, *k* = 10, *β* = 0.5, *T* = 20 and *τ* = 4. In each panel, reducing transmission means reducing *β* by half, distancing means reducing *k* (the number of people in proximity) by half, and “strict bubbles” means ensuring that attendees contact only *k* individuals over the whole event rather than mixing with others outside their bubble. *Top*: no mixing (*τ* = *T*); the horizontal axis is the total event duration in hours. *Middle*: Mixing occurs every 4 hours. *Bottom*: A setting with a 10x lower propensity for transmission (*β* = 0.05). Here, transmission never “saturates” because 1 − *e*^−*βτ*^ remains small enough that it is approximately *βτ*, which is small.

Fig. 1 (right) shows how *eR* changes with respect to time for some different settings and with different interventions. The top panel shows the impact on *eR* for events without mixing. When the event’s duration is short, reducing transmission (for example with masks and barriers) and ensuring distancing have similar impacts, but when the duration is long, reducing transmission has much less impact than distancing. As the middle panel shows, at events where individuals mix, strict bubbles can be much more effective than either distancing or reducing the transmission rate, and distancing out-performs reducing transmission. However, when the baseline transmission rate is very low (bottom panel), distancing and reducing transmission are better than strict bubbles. Here, contacting three different groups of 10 people for one hour and contacting a single group of 10 people for three hours will each result in the same (low) average number of new infections. We refer to events like these as “linear” events: the expected number of new infections depends linearly on the number of contacts and the duration. In contrast, if the transmission rate is high enough that exposure of length *τ* can lead to a substantial fraction of the first group of *k* people becoming infected, then it is far preferable not to move to a new group of *k* people when that hour ends. We refer to such events as “saturating”.

In order to use our model to make recommendations for the planning of specific events it is necessary to estimate the transmission rate *β*, since it is not directly observable. We obtained reports of outbreaks at a range of events including parties, meals, nightclubs and restaurants. For each, there was sufficient information to estimate *eR* and the duration *T*. For example, 52 of 60 singers became infected after a choir rehearsal in Washington (*12*); 5 of 39 passengers were infected in China when a man took a 2-hour bus ride without a mask, whereas none of 14 passengers on his next, 50-minute, bus journey were infected when he wore a mask (*21*). 19 people were infected by a single individual in a nightclub outbreak (*2*). See Supplementary Information for the complete list.

We used images of similar events to estimate *k*, under the assumption that two people were in contact if they were within two meters of each other. The mixing time *τ* was estimated given the nature of the event. All parameters *k, τ, eR* were sampled widely to accommodate the considerable uncertainty in these quantities. Using Eq. 1, we then obtained samples of *β* Fig. 2. Transmission rates range from 0.02-0.05 transmissions per hour (from household studies, a funeral) to 0.5-0.6 transmissions per hour (choir, party, lunch), with events involving speaking, singing and eating (parties, meals) generally higher than those without. We also estimate the turnover (1−*τ*/*T*) and saturation (1−*e*^−*βT*^); broadly, saturating events with high turnover have the highest *eR* and therefore are the highest risk Fig. 2. We note that these are likely over-estimates of *β* due to the fact that exposures that did not lead to transmission are not included and that larger outbreaks are more likely to be widely reported (see Supplementary Information). However, it is precisely these larger outbreaks that are of most concern.

**Figure 2:**
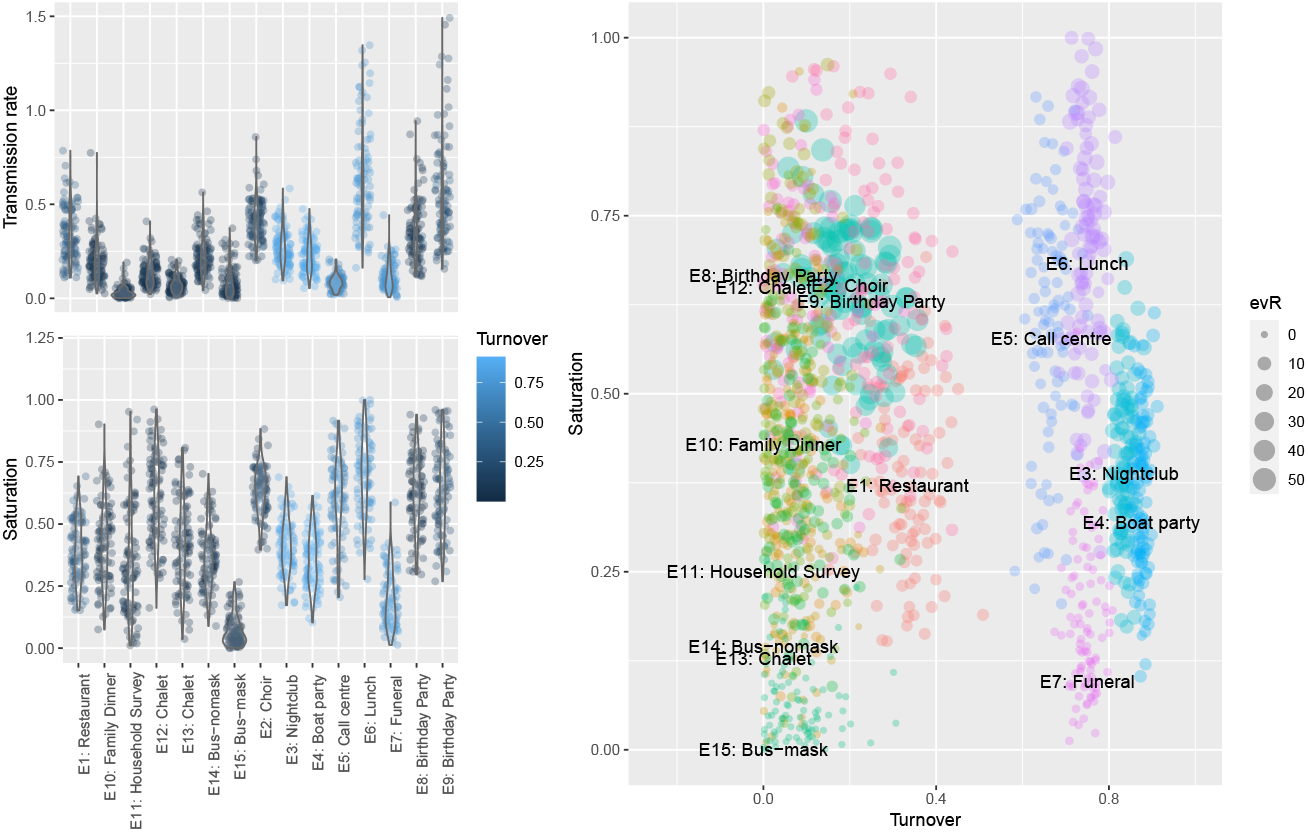
Transmission rate and saturation vary over reported events with transmission rates in the range 0.02-0.5 transmissions per hour (given contact). Transmission rates are highest for events involving sharing meals, singing and speaking (presumably at volume, though we do not have this information). Among the events we described, the choir, birthday parties, call centre and lunch are the most “saturated”. The re-sampled parameters (100 per outbreak) yield variable *eR* but overall, saturating events with high turnover present the highest numbers of infections per infectious attendee.

These numbers can be used to explore the impact of interventions in new settings. For example, consider a crowded indoor event such as a sporting event, crowded conference evening, rally or rock concert, where *k* would be about 15, and the duration *T* approximately three hours. We would expect some mixing (*τ* of one hour) and an indoor transmission rate in the range of 0.2−0.4 per hour. This gives *eR* in the range 4 −14; if *β* = 0.4, the event is 70% saturated (1−*e*^−*βT*^ = 0.70) and *eR* = 14. Spacing people so that *k* is halved reduces *eR* from 14 to 7; halving *β* reduces *eR* to 8 and strict bubbles of 15 reduce *eR* 10. Reducing both transmission and density reduces *eR* to 4. In contrast, consider elementary and high schools. In elementary schools students remain in the same class group throughout the day, and in high schools each class has a new mix of students. For one week of high school with *T* = 24, *τ* = 3, *k* = 10, and *β* = 0.3 (based on similar cases in our data), *eR* is 47. Halving *β* reduces *eR* to 28 and halving *k* reduces *eR* to 24. But if we structure into fixed classes as in elementary schools, with *τ* = *T*, this reduces *eR* to 10, which is more than twice as effective as the other measures.

We propose that organizers, workplaces, businesses and so on seek to determine if their setting is likely to be linear or saturating, and whether people mix strongly or remain in small groups (or “bubbles”) – see Fig. 3. In all events, interventions that increase distancing (reducing *k*) are effective. In events that are already static, the relative importance of reducing transmission (reducing *β*) is much greater in the linear setting. For events where there is mixing, bubbling (reducing *τ*) is an extremely powerful intervention in the saturating case, but is less significant in the linear case.

**Figure 3:**
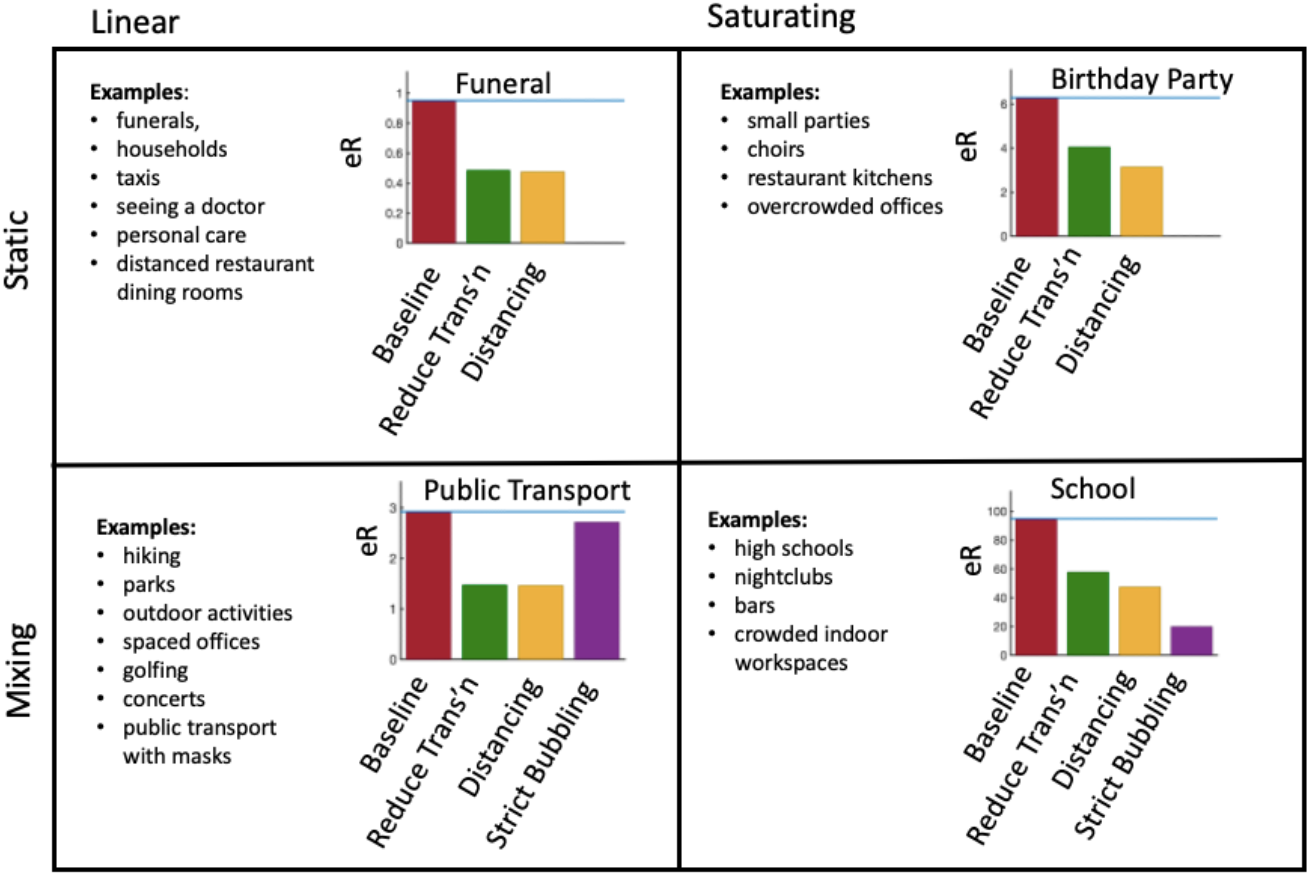
Four different kinds of events depending on whether they are *linear* (low transmission probability) or *saturating* (high transmission probability); and whether they are *static* (same contacts for whole event) or *dynamic* (high turnover of contacts). We select representative parameters for each type of event, determine the number of new infections, and show how the three interventions effect this number. Interventions are reducing transmission (halving *β*), introducing distancing (halving *k*), and strict bubbling (setting *τ* = *T*). The parameters used for the plots are: *Funeral*: *k* = 10, *τ* = 2, *T* = 2, *β* = 0.05, *Birthday Party*: *k* = 9, *τ* = 3, *T* = 3, *β* = 0.05, *Public Transport*: *k* = 15, *τ* = 1, *T* = 4, *β* = 0.05, *School*: *k* = 20, *τ* = 3, *T* = 24, *β* = 0.3.

Saturating situations may not only make reducing transmission challenging, they may also make it difficult to estimate the effectiveness of masks and other physical barriers to transmission. This is because in saturating settings, even an intervention that halves the transmission rate may not have much impact on the number of infections. This effect may help to explain the variable evidence for the benefits of masks in reducing transmission, with some studies showing no benefit (*16*), while the overall picture shows significant benefit in some cases (*5*). In contrast, the evidence that transmission is impacted by physical distance is quite strong. Distancing of one metre or more significantly reduces transmission, and greater distances reduce it further (*5*). *Strict* bubbling can be effective, and has the added advantage that contact tracing is made easier when individuals have fewer contacts and can identify them, but strict bubbles are hard to maintain over time due to social, family and workplace activities.

Our fundamental relationship focuses on *eR* which can be seen as an average over a number of heterogeneities, including variation in individual infectiousness. It identifies the potential for “superspreading events”, particularly saturated and highly mixing events, which can have very high *eR*. The total number of infections associated with an activity will depend not only on *eR*, but on the frequency of the event, the total attendance, and the prevalence of the disease in the population. For example, while *eR* for a 30-minute bus ride is likely to be low, transit authorities must make decisions that account for the number of transit users, the frequency with which they take transit, as well as COVID-19 prevalence. With both benefits and expected transmissions depending on the number of people engaged in an activity or event, societies must decide which events and activities have an acceptable COVID-19 cost-benefit balance. Decision-makers must also consider ongoing community transmission subsequent to events; individuals attending one type of event may be likely to attend others, amplifying the effects. Dynamic transmission models can help explore the impact of superspreading events in the context of broader transmission (*15*).

Complex settings such as universities have a population engaged in a series of “events”: classes, movement between classes, dining halls, dormitories and transportation to campus. While we would suggest that within a class, assigned seating, distancing and mask use will likely combine to reduce transmission considerably, close contact in dormitories and dining halls could still result in transmission. Our framework could help design measures targeted to each activity, but these would likely need to be supplemented with rapid contact tracing and case finding. It is essential to support exposed individuals to be able to isolate themselves without suffering economic, social and educational consequences.

A range of new outbreak settings will likely be reported as more activities re-open (*18, 33*). The largest outbreaks reported to date have naturally included cases arising over many days, and have taken place in long-term care facilities (*24*), meat and poultry-packing facilities (*8*), correctional facilities (*1*) and other high-transmission environments (*9, 28*). These may be saturating, mixing environments, which in our framework helps to explain high case volumes, although we did not find that the call centre (*25*) was saturating. These settings have a fixed population and long durations, whether individuals are present full time (patients and inmates) or for full working days (staff). In a closed setting with a fixed population, if the event’s duration is defined to be the duration of infectiousness, event R is the classic “basic reproduction number”, *R*0 (the expected number of new infections an individual is expected to create in a fully susceptible population). The eventual size of an outbreak in a closed setting depends on *R*0 and on the interventions taken to stop transmission (*3*).

The possibility that some individuals are infectious but never develop symptoms (*19*) could mean that they attend a setting for a period *T* of many days, creating a “saturating” setting even if the transmission rate is low. In this case, mask use and physical barriers to transmission may be ineffective, physical distancing is likely to be more effective and strict bubbling the best. In addition to the risks posed by asymptomatic individuals (who may after all not be as infectious as others (*22*)), even for those who eventually develop symptoms it has been estimated that over 40% of transmission occurs before symptom onset (*29*), over a period of a few days. In our framework, with the transmission rates we have acquired from reported short outbreaks, a time period of several days places some activities firmly in the “saturating” mode.

While we do not currently have data to determine the relative COVID-19 risks for most activities, we should begin collecting this information prospectively, noting *k*, the extent of mixing, outbreaks’ duration and location, and how many individuals are infected by a single index case in a given setting. Centres for disease control who maintain contact tracing programs, together with workplace, venue and facility staff, could collate these data. If digital contact tracing apps are introduced (*4*), these could provide extremely rich data on the parameters in our framework, on *eR* itself, and on the settings in which exposure occurred but infection did not. Our framework, together with these data, can then inform what the most effective, feasible measures are for particular settings.

## Data Availability

All data are publicly available at the links given in the text.

## Acknowledgments

PT, CC and MY were supported by Natural Science and Engineering Research Council (Canada) Discovery Grants (RGPIN-2019-06911 and RGPIN-2019-06624). CC receives funding from the Federal Government of Canada’s Canada 150 Research Chair program and from Genome BC (COV-142).

## Supplementary Information

### Materials and Methods

#### Reported outbreaks

Our starting point for fitting our model to COVID-19 outbreak data was a database of reported clusters in the scientific literature and news media (*20*). From the more than 100 outbreaks described there we selected a small number of incidents where there were enough details reported for us to estimate our parameters. Most reported outbreaks list the total number of infected individuals, including two or three generations of infection, and individuals who were not at the event in question. Since our *eR* is defined to be the number of new infections directly caused by one infected individual at the event in question, we selected outbreaks where: (1) there was likely only one infected individual initially at the event, and (2) where there was an estimate of how many people were directly infected by this individual at the event, or there was information about the timing of the appearance of symptoms in all infected cases. This meant that we could estimate *eR*, using information about the time interval between infection and the expression of symptoms. For each event we selected maximal and minimal values of *eR* that were consistent with the reported data.

*T, τ*, and *k* were estimated using the description of the events where the outbreaks occurred. Often *T* was reported, but otherwise we picked a reasonable number for events of that type. For example, we selected *T* = 2 hours for a funeral. There was no specific data available for *τ* and *k* for any of the outbreaks. *τ* was guessed using what was known about the type of event. For example, in a choir people typically stand in the same place for most but not all of the duration of the practice, and so we set *τ* = 2, *T* = 2.5. As for *eR*, we chose a range of values for each *k* based on the principle that transmission can occur when individuals are within two meters of each other. In many cases our estimates were just informed by our own experiences of such events. For others, we were able to estimate ranges for *k* using photographs of similar events; see below for details.

#### Uncertainty

We took the following approach to incorporating uncertainty in the parameters from our outbreaks. Given our range for *k* we sampled *k* from a normal distribution whose mean is the midpoint of the range, and whose standard deviation is 1/4 the range (so that 95% of the samples lie within the estimated lower and upper values). We took the same approach for *eR* (using our estimates of upper and lower values and using a normal distribution to sample primarily within that range). For *τ*, we interpreted our estimated *τ* above as a mean 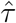 and sampled *τ* from a normal distribution with standard deviation 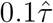. In outbreaks with little to no mixing 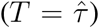, we reflected the samples with the mapping *τ*_*r*_ = *T* − |*T* − *τ*|, where *τ* is the sample from 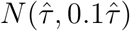, and *τ*_*r*_ is reflected so that the re-sampled values are always less than the total time *T*.

As stated in the main text, for each choice of the parameters *k, T, τ* and *β*, according to the model the expected number of new infections is

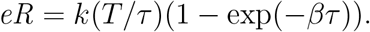

But given a set of parameters, the actual number of new infections *X* is a binomial random variable with parameters *p* = (1−exp(−*βτ*)) and *n* = *k*_*e*_ = *k*(*T*/*τ*). We used a standard Bayesian framework to determine a probability distribution for *p* and hence *β*. (See (*10, Ch. 2*) for an exposition of this case.) Let the observed number of new infections be *X*, where

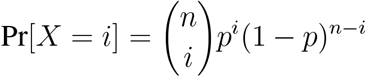

Given that we observe *X* = *eR*, and assuming a uniform prior on *p*, this gives the likelihood for a given value of *p* proportional to

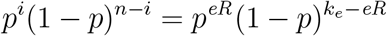

which is a Beta distribution with shape parameters (*α, β*) = (*eR* + 1, *k*_*e*_ −*eR* + 1). Once *p* is sampled from this distribution, *β* is then given by

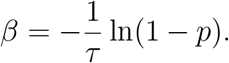

For each event we generated the points in the plot in Figure 2 by (1) selecting *k, eR, T, τ* at random from the distributions described above; (2) generating a value of *p* from the above Beta distribution and (3) inverting to obtain a sample of the transmission rate *β*, which we then show in Fig. 3 and use to explore the impacts of interventions in different events.

Despite our attempts to account for the many sources of uncertainty in our data when we obtain our posterior distributions for *β*, there is an important source of bias we could not account for. By definition, an outbreak could only enter our study if there was some transmission. Likewise, the likelihood of an outbreak being noticed and entered into a database probably increases with the number of infected individuals. This means that we are observing unusually large *eR* for the events that occur, and so are overestimating *β*. It might be possible to adjust for this effect using carefully collected datasets (*9*) but fundamentally this would require knowledge of exposures that did not lead to infections, and this is seldom collected systematically.

### Approximating *k* with images

An important parameter in our model is *k*, the number of people within transmission range of an infected individual. We fixed the transmission range to two metres, and sought to estimate *k* for various events using images of similar events obtained via Google image search. After one person in the image was chosen, we used the fact that average shoulder width is approximately 40cm (*31*) to estimate what 2 metres corresponded to in the image. We then counted the number of people within that range of the given person. We repeated the same process for another person on the image if possible, and for several images. After several such counts, we could obtain a range of values for *k*, which we show in the following table.

| location type | image links | lower bound $k$ | upper bound $k$ |
| --- | --- | --- | --- |
| nightclub | 1, 2, 3 | 20 | 35 |
| bar | 1, 2, 3 | 15 | 25 |
| restaurant | 1, 2, 3 | 10 | 15 |
| conference | 1, 2, 3 | 5 | 15 |
| gym | 1, 2 | 5 | 10 |
| church | 1, 2 | 15 | 25 |
| funeral | 1, 2 | 10 | 25 |
| lecture | 1, 2 | 5 | 15 |
| hallway | 1, 2 | 5 | 25 |
| swimming pool | 1, 2 | 5 | 10 |
| bus | 1, 2 | 5 | 25 |
| choir | 1, 2 | 20 | 30 |

### Stochastic mixing model

We chose an oversimplified model of interaction in dynamic environments in order to capture the essential ideas and have simple closed-form solutions. But the key features of our analysis remain true for more realistic models of mixing. Here we demonstrate such a model and also illustrate how these models can be simulated stochastically.

First we describe a probabilistic simulation of a static situation, which we show in the top plot of Fig. S1. We imagine a single infected individual in the presence of *k* = 10 susceptible individuals, for *T* = 20 hours, with a transmission rate of *β* = 0.5, which leads to a saturating situation. We simulate transmission by choosing a small *δt* = 0.01 hours and for each successive time interval of that length infecting each of the remaining susceptible individuals with probability *δtβ*. The plot shows the number of newly infected individuals as a function of time in the baseline case and the two interventions where *β* is halved and *k* is halved. Results are similar to those for the deterministic simulation in Fig. 1 in the main text.

For a dynamic simulation with a more complicated mixing model than in the main paper we imagine a single infected individual walking past a long line of susceptible individuals and interacting with them in turn. The individuals are spaced so that the infected individual is always within range of *k* initially susceptible individuals. The person walks at a speed so that every *τ* time units there is a complete replacement of the *k* people in range. However, susceptible individuals enter and leave the walker’s range one by one. As above, transmission occurs at rate *β* for every susceptible individual in range of the infected individual. The infected individual walks past the line for total duration *T*. The simulation is performed in the same way as in the static case, whilst keeping track of which individuals are in range of the walker. The middle and bottom plots of Fig. S1 show the results for the same parameter choices as in Fig. 1, and we see that the relative performance of the different interventions remains unchanged.

### Variability in transmission

There are several ways in which the transmission of COVID-19 is likely more complex than indicated in our model.

As one example, if a threshold viral inoculation is required for infection, and if short exposures can be under that threshold, then multiple very short exposures could be preferable to fewer longer exposures. This would change our conclusions about mixing, and would result in a “sub-linear” regime in place of the linear one at low transmission rates. We have also not included a possible infectious dose-severity effect; if higher viral inoculations lead to more severe disease, then in saturating settings it is crucial not only to reduce mixing but also to reduce viral dose (for example with masks and other barriers).

Furthermore, we did not explicitly incorporate factors that affect the rate of transmission from an individual. These include the individual’s severity and viral load (*22*), time since infection, droplet production, and behaviour, among others. In Eq. 1, we could represent these factors during the course of infection using an infection-age-dependent *β*_*i*_(*a*) (where *a* is the time since infection and *i* denotes a “host type”). Naturally we do not know at which stage an infectious individual may attend an event, and we do not fully understanding variability in infectiousness among individuals. If we knew the infectivity profile over time, and denote the infectious profile in a host of type *i* (in the relevant setting), we could write the probability of transmission after a time *τ* given a host of type *i* a time *a* after infection as

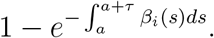

In this case, what was *βτ* is now 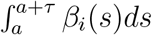. When *τ* is much shorter than the duration of infectiousness, as in all of our examples, the integral is well approximated by *β*_*i*_*τ*. The overall framework of linear, saturating, mixed or static events remains the same; the transmission rate *β* depends on the event and the infectious individual, and in our exploration of reported outbreaks we have explored *β* for particular event-person pairs.

### Outbreak Details

Here we briefly describe each of the outbreaks shown in Fig. 1 along with how we obtain our range of values for our parameters.

#### E1: Restaurant

This outbreak was traced to a single infected individual eating dinner at a restaurant in Gaungzhou, China (*23*). A range for *k* was estimated by looking at the restaurant seating plan, but also allowing a larger *k* for the hypothesized transmission by air conditioning in this setting. Nine people at the restaurant at that time were eventually infected; taking into account certain and probable secondary infections put *eR* between 5 and 7.

**Figure S1:**
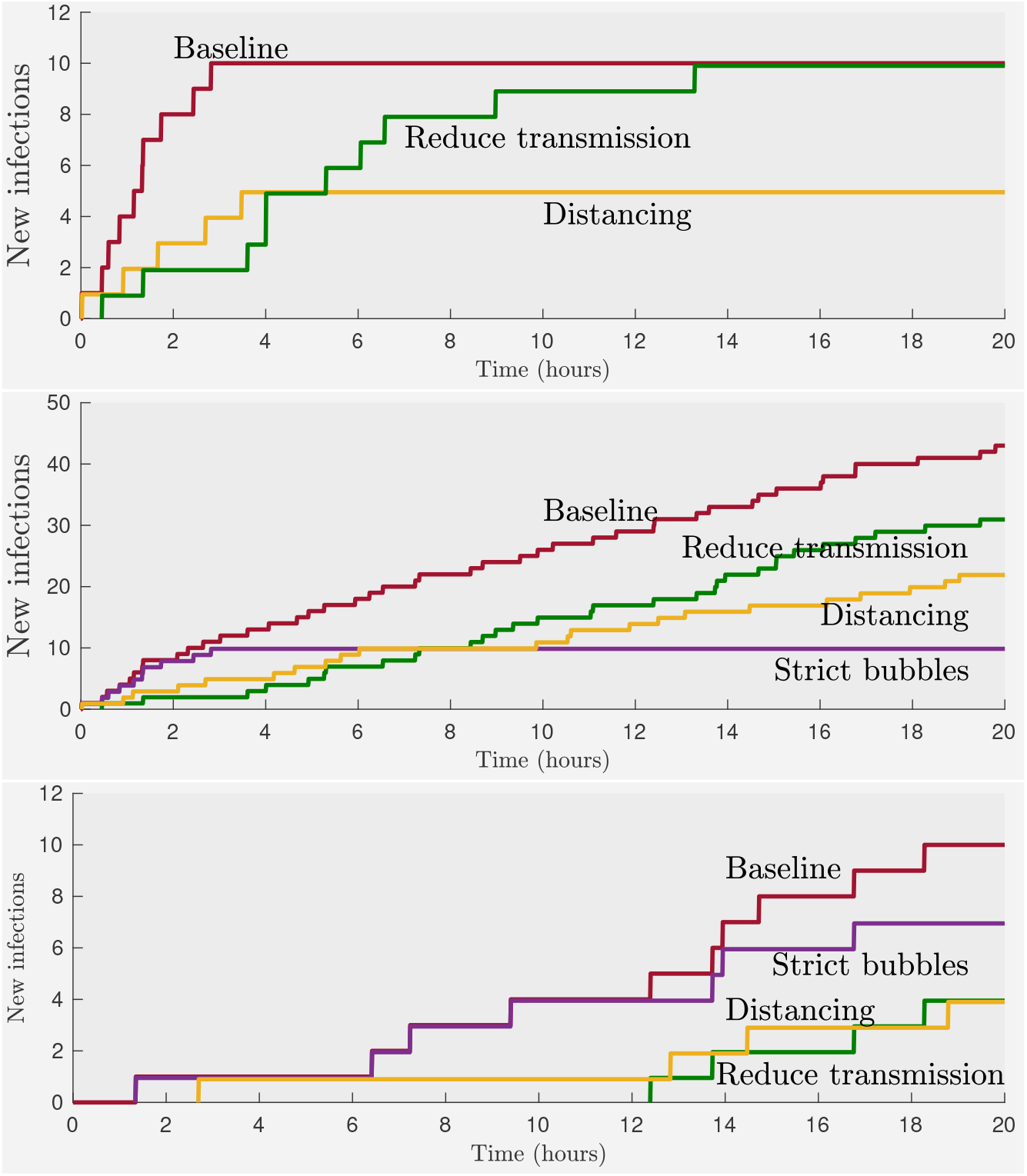
Results of the same simulations as in Fig. 1 but with the stochastic model of interaction.

#### E2: Choir

In this now famous case a single infected individual led to the infection of probably 52 of 60 other individuals in a choir (*12*). The report determined that all of the transmission from the index case occurred in a single choir practice. We followed this assumption, even though we think it possible that some transmission occurred asymptomatically during the practice a week before. We assumed that all people present were potential contacts (*k* between 55 and 60) and that *τ* = 2, *T* = 2.5. Certainly it is possible that all 52 cases were infected at the practice, but some of the reported cases were probable and not confirmed, and we also cannot rule out secondary infection if some choir members socialized together before or after the practice. So we chose a range of *eR* between 30 and 52.

#### E3: Nightclub

According to a news report 19 people were infected by a single infected individual (*2*). There was not any data to determine how many of these infections were secondary, but we estimated a range of 10 to 16 for *eR*. For the *k* range we used our image data for nightclubs; see above.

#### E4: Boat party

In this event it is believed that a single taxi driver who had transported passengers from Wuhan led to the infection of 10 others at a party on a boat with approximately 90 guests total (*32*). Based on excluding possible secondary cases we chose *eR* to run between 5 and 10. The approximation for *k* was chosen based on the ranges of *k* for restaurants and nightclubs cases as being similar settings.

#### E5: Call center

This synopsis article describes the outbreak at a call center (*25*). Out of 1143 people tested 97 were confirmed to have the disease, and in particular, 94 worked in a call center on the 11th floor of the building, mostly on the same side. The first case-patient (with symptoms onset) worked on the 10th floor and never went to the 11th one. The second case-patient worked at that call center on the 11th floor. *k* was computed using the detailed floor plan provided by the article. A range for *eR* was estimated using the epidemic curve which was provided in the paper.

#### E6: Lunch

According to this news article (*26*), at least 60 out of 850 members of Rio de Janeiro country club were confirmed to have the virus. The source mentioned the lunch at the mansion, where more than half of the approximately 70 guests tested positive. Assuming that there were many secondary infections we chose *eR* to range from 7 to 18. Since we didn’t know the arrangement of seating at the lunch we gave *k* a particularly wide range, from 5 to 20. We assumed that lunch lasted *T* = 2 hours with a lot of mixing *τ* = 0.5.

#### E7: Funeral

In this report (*11*), 4 attendees of a funeral were tested positive for the virus. The transmission happened at the meal and/or following funeral lasting 3 and 2 hours respectively. Based on descriptions of the events and the timings of symptom onsets, we believe that there was only one actual transmission at the funeral. *k* was chosen to be between 5 and 15.

#### E8: Birthday party I

In the same report (*11*), three days after the funeral, a birthday party took place with 10 attendees including the index case from the funeral. 7 out of 9 developed the virus after several days. The article reports close contacts among the attendees leading to *k* = 9. Based on the detailed contact tracing, *eR* was estimated to be between 5 and 7.

#### E9: Birthday party II

According to this news report (*13*), 7 out of 25 birthday party attendees are believed to have caught the virus from a single infected individual. (Eventually a total 18 family members were confirmed infected.) We chose a wide range for *k*, lacking data about how the party was organized.

#### E10: Family Dinner

The reference (*14*) describes in detail an outbreak brought to Nanjing, China by a traveller who had passed through Wuhan. Ten other people were infected in a variety of events, but we focused on one family dinner where 3 people were infected out of the 7 guests. We chose *T* = *τ* = 3 hours.

#### E11: Household Survey

This reference (*30*) is different from the others in that it surveys transmission in the households of 85 patients that were infected with COVID-19. Household sizes ranged in size from 1 to 7 other people (giving us our range of *k*) and in the vast majority of cases at most 2 other people were infected, giving an *eR* range running from 0 to 2. We assumed that individuals in households would spend 8 hours a day together, and that they would have two days of opportunity to transmit the virus before they practiced preventative measures, giving *T* = *τ* = 16 hours.

#### E12 and E13: Chalet

This outbreak occurred in the French Alps where an infectious British tourist stayed at two separate chalets (*6*). The data is sufficiently detailed that we could estimate parameters for each of the chalet visits. At the first chalet there were *k* = 10 other adult guests, 9 of which were eventually infected. Taking into account of secondary infections we put the *eR* range as running from 4 to 9. At a second night at a different chalet 3 out of 5 others were eventually infected; we chose *k* = 5 and *eR* between 1 and 3. We assumed *T* = *τ* = 8 hours for both nights.

#### E14 and E15: Bus trips

An infected individual from Chongqing, China took two bus trips in quick succession, the first with a face mask, and the second without (*21*). Five people were infected on the first bus trip and none were infected on the second. We estimated ranges for *k* in each case by looking at plans of buses, and using the fact that the second bus was a minibus. The durations of the bus trips were recorded quite accurately and in each case we set *T* = *τ*.

